# Fluid administration guided by inferior vena cava ultrasound before spinal anaesthesia may reduce post procedural hypotension rate

**DOI:** 10.1101/2021.06.20.21258944

**Authors:** Mathieu Favre, Samuele Ceruti, Maira Biggiogero, Michele Musiari, Andrea Glotta, Isabella Gimigliano, José Aguirre, Alain Borgeat, Andrea Saporito

## Abstract

**PURPOSE:** This study was conducted to estimate the incidence of hypotension after spinal anaesthesia after inferior vena cava ultrasound (IVCUS) guided volaemic optimization compared with a control group in patients undergoing elective surgery. According to ESICM guidelines, hypotension was defined as two systolic arterial pressure (SAP) measurements < 80 mmHg and / or a mean arterial pressure (MAP) < 60 mmHg, or a drop in SAP of more than 50 mmHg or more than 25% from baseline, or a decrease in MAP by more than 30% from baseline and / or clinical signs/symptoms of inadequate perfusion.

**MATERIALS AND METHODS:** From May 2014 to February 2019, a prospective, controlled, randomised, three-arm, parallel-group trial was performed in our tertiary hospital. In the IVCUS group (I, 132 patients) and passive leg raising test group (L, 148 patients), a pre-anaesthesia volume optimization was achieved following a fluid response protocol. In control group (C, 149 patients), no specific intervention was performed.

**RESULTS:** 474 patients were collected. In group I, hypotension rate was 35%. In group L hypotension rate was 44%. In group C hypotension rate was 46%. An 11% reduction rate in hypotension (95% CI -1 to -24%, P=0.047) was observed between the group I and the group C. A 2% reduction rate in hypotension (95% CI -3 to -5%, P=0.428) was observed between group L and the group C. Total fluid amount administered was greater in the I group I than in the group C (593 ml versus 453 ml, P=0.015) and greater in the group L than the group C (511 ml versus 453 ml, P=0.11).

**CONCLUSION:** IVCUS guided fluid optimisation decrease the incidence of arterial hypotension after spinal anaesthesia.

## INTRODUCTION

Spinal Anaesthesia (SA) is safe and used for different surgeries [1]. Arterial hypotension is one of the most frequent adverse events [2 - 6]. Transient hypotensive episodes are generally well tolerated in patients without particular comorbidities [7, 8]. However, these episodes may lead to major complications in elderly emergency situations and cardiovascular high-risk patients [2, 4, 9] or those who take medications like beta blockers [2], angiotensin converting enzyme inhibitors, selective serotonin recapture inhibitors or mono-amine oxidase inhibitors [10]. Arterial hypotension is usually prevented by empirical fluid administration [3, 4, 9, 11], without using specific tools to evaluate patient’s hemodynamic status. In patients with pre-existing disease this practice may increase the risk of volume overload, further impairing cardiac and lung function, potentially leading to congestive heart failure and/or pulmonary oedema [4, 11].

Currently, two main non-invasive tests are performed to predict and identify fluid responsive patients in a spontaneous breathing situation. The Passive Leg Raising Test (PLRT) consists in raising passively the patient’s legs to increase venous return and therefore cardiac output [12]. Until now, PLRT is considered to be one of best methods [11 - 13] to predict fluid responsiveness in spontaneous breathing patients, measuring changing in stroke volume (SV) by echocardiography or other non-invasive methods. Inferior Vena Cava Ultrasound (IVCUS) is another useful test that analyses IVC’s variability during spontaneous breathing activity [14, 15]. Several studies previously conducted, demonstrated advantages and limitations of these methods in spontaneous breathing patients [15 - 19], but there is no test guided volaemic fluid response protocol in the specific setting before SA [18].

Previous publication of the *ProCRHYSA trial* [20, 21] aimed to determine whether IVCUS and PLRT were effective in guiding fluid therapy both to reduce the rate of hypotension and the needed amount of volume in non-critical patients undergoing SA for elective surgical procedures. To answer these questions, we present the final results of the ProCRHYSA trial.

## METHODS

### Design

This was a prospective, controlled, randomised, three-arms, parallel-group trial of consecutive patients undergoing elective surgery under SA treated in our hospital. This trial was registered on ClinicalTrials.gov and was approved by local Ethical Committee. Written informed consent was obtained from patients prior to randomization into three parallel groups, with an allocation ratio of 1:1:1. Using an electronic software, group assignment was performed by an independent investigator not involved in patient treatment.

Inclusion criteria were adult patients of both sexes, evaluated as American Society of Anaesthesiology (ASA) risk class I to III [22], undergoing elective intervention under SA. Exclusion criteria were patients requiring invasive blood pressure monitoring, patients affected by pre-procedural hypotension (systolic arterial pressure less than 90 mmHg and/or mean arterial pressure less than 60 mmHg), patients with unilateral anaesthetic block, patients requiring any form of assisted ventilation prior or during the surgical intervention and patients unable to give informed consent [23]. Contraindications to perform SA were previous back surgery in the lumbar region, clinical high-risk conditions like thrombocytopenia (<50 G/L) or coagulation abnormalities. Obstetrical patients were also excluded [23].

Arterial hypotension after SA was defined as two measurements of systolic arterial pressure inferior to 80 mmHg and/or a MAP inferior to 60 mmHg [23], or a fall in systolic arterial blood pressure of either more than 50 mmHg or more than 25% from the baseline value, or a reduction in mean arterial pressure of more than 30% from the baseline value and/or clinical signs/symptoms of inadequate perfusion [20, 21]. All these parameters were collected in a specific software, in order to automatically identify hypotensive patients.

### Setting

The study started upon the arrival in the operating block (time 0) and ended 30 minutes after completion of SA. Three phases were identified; the *pre-anaesthetic phase* started from time 0 to the beginning of SA, the *anaesthetic phase* corresponded to the performance of the SA and the *postanaesthetic phase* from the end of anaesthesia to the next 30 minutes (Fig. 1), which corresponded to the end of the patient’s study period.

**FIGURE 1:**
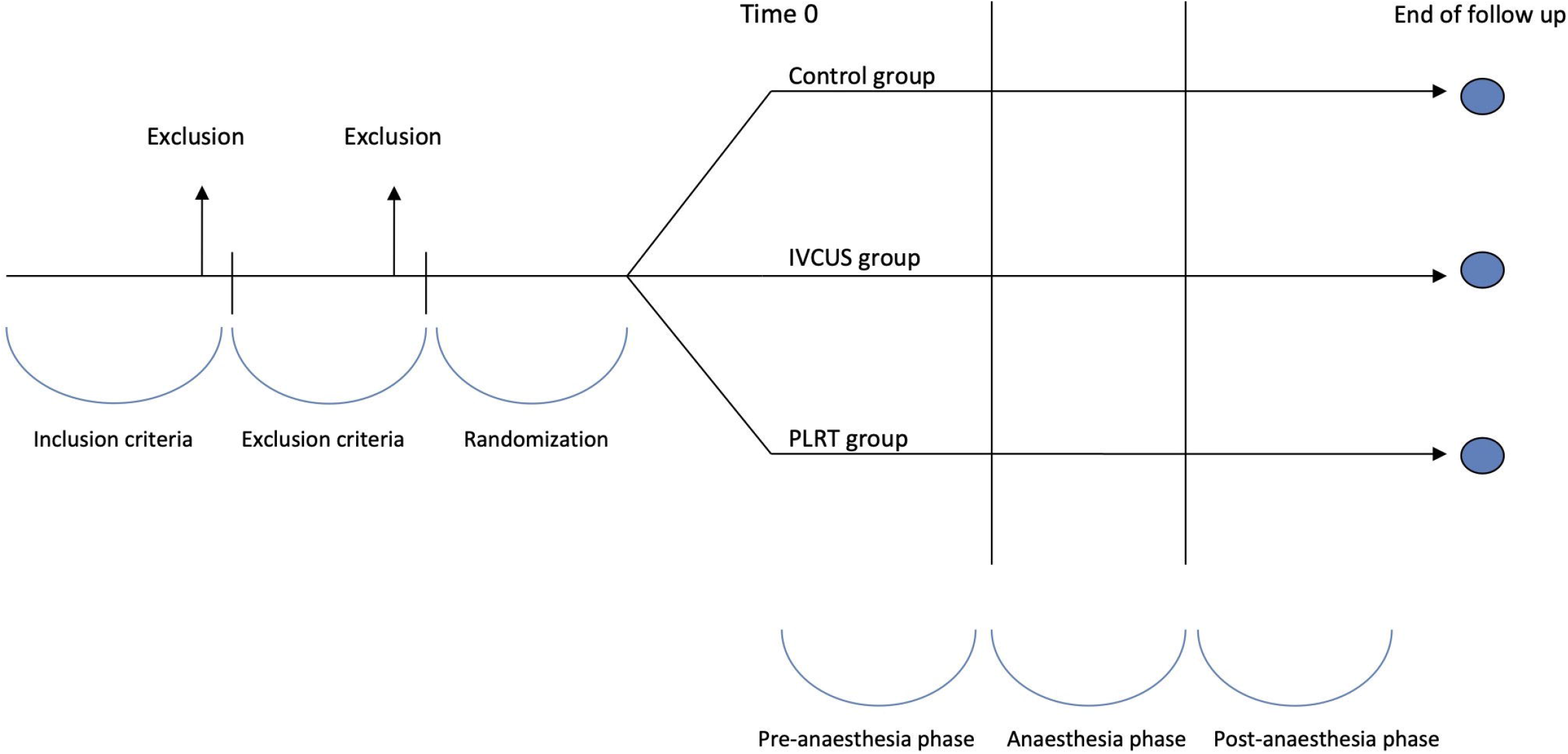
Study design. During *pre-anaesthesia phase*, fluid status of patients enrolled in IVCUS and PLRT arms had be optimised following the relative study procedure. At *anaesthesia phase*, patients from these two groups were not more fluid-responders according to relative procedures. Outcomes (hypotension rate, fluid and drugs management) were verified in *post-anaesthesia phase*.

#### Control group

In the induction room, patients were monitored with a continuous three lead ECG, pulse oximetry and non-invasive blood pressure measurement with cuff inflating every minute for continuous hemodynamic monitoring. An IV line was obtained by inserting an 18 gauge cannula. No intervention was performed before SA, with the exception of a minimal filling of crystalloids (pre-load) according to the duration of the preparatory procedure and the time of anaesthesia execution. SA was subsequently performed in a lateral decubitus following a standardised procedure: a dose of 12-18 mg hyperbaric bupivacaine 0.5% was administrated intrathecal at median L_3_-L_4_ level by using a 27 gauge bevelled pencil point, cranially orientated spinal needle (BBraun Medical SA, Melsungen, Germany) on the basis of the type of surgical procedure and patient’s morphotype. Immediately after the injection, patients laid supine for 30 minutes with another minimum amount of crystalloids (co-load) administered according to usual clinical practice. A nurse not involved in the trial was assigned to assess the sensory block extension with cold test for the targeted level block [23].

Post SA hypotension was treated with crystalloid replacement (250 ml bolus) and vasoactive drugs (Phenylephrine 100 ug IV), until hemodynamic stability was reached. All clinical and pharmacological data were collected.

#### IVCUS group

Patients allocated into this intervention group were monitored in the same way as those allocated in the control group, with an additional fluid status assessment by IVC ultrasonography (Philips Sparq ultrasound) prior to SA (Fig. 1). As previously described [20], the subcostal echocardiographic window was evaluated to perform the IVC measurement in spontaneous breathing patients. The IVC collapsibility index (IVC-CI) is intended as the difference between the IVC measured at its maximum diameter (final expiration) and minimum diameter (final inspiration) divided by the maximum diameter. According to the great majority of authors [14, 15, 17, 24, 25] and to our published protocol [20], IVC-CI greater than 36% [20] was chosen and used as cut-off level. As there is no published consensus concerning the amount and type of fluid filling [11], rapid administration of 250 ml of crystalloid infusion in 10 min was considered adequate and safe.

According to IVC-CI, patient’s hemodynamic status was re-assessed after each refilling: if the patient resulted as *responsive*, it was refilled once again; on the other hand, when the patient became *unresponsive*, SA was performed as described for the control group. Each time, only 1 IVC-CI was calculated. Follow-up was identical compared with control group. Thirteen certified physicians performed the ultrasounds analysis during the entire study period.

#### PLRT group

Patients allocated in this group were monitored in the same way as in the control group; PRLT and fluid administration was performed prior to SA until the test was normalised (Fig. 1). PLRT had been shown to be a highly accurate method to predict fluid responsiveness in spontaneously breathing critically ill patients [12]. Active leg elevation exerted an orthosympathetic reflex which can increase cardiac output; passive lower limb test had the advantage of mobilizing lower limb venous blood (about 300-500 ml) without activating the orthosympathetic reflex, making possible to quantify the clinical response after a bolus of fluids.

An etCO_2_ measurement throughout continuous CO_2_ sampling through nasal cannula with patients in semi-recumbent supine position at 45° was also performed, before and after the PLRT; before the test, patients waited 5 minutes in a semi-recumbent supine position to reach a phase of hemodynamic and respiratory balance and general stability, also to reduce anxiety and stress. During the test, an etCO_2_ increasing more than 12% from baseline was interpreted as *fluid-responsive* [12]. Prior administration of SA, patients were subsequently stratified as *responsive* or *unresponsive* and managed according to the same protocol as described for the IVCUS group; after each refilling, patient’s hemodynamic status was re-assessed through etCO_2_ variation. SA was performed once unresponsiveness was reached. Follow-up was identical compared with other groups.

### Outcomes

*Primary outcome* was the hypotension rate after SA following fluid optimization therapy guided by IVCUS and PLRT test or no intervention. *Secondary outcomes* were an analysis on fluid amount administered, on vasoactive drug amount used and time needed to achieve the whole anaesthetic procedure.

### Statistical analysis

From previous studies we estimated the incidence of arterial hypotension after SA to be of about 50% in the control group [20]. In order to have a significant reduction of 10% between IVCUS group and control group, with a power of 80% and a significance level of 0.05 (one-tailed z-test) a sample size of 130 patients per group was calculated. Calculating a drop out of 13-15%, due to technical difficulties, a sample size of about 150 patients per group was desired. Descriptive statistics of frequency was performed; data were reported as number (percentage). Data distribution was reported as mean ±SD if normally distributed, otherwise as median (IQR); all data distribution was verified by Kolmogorov-Smirnov test. Differences between continuous variables were studied by t-test; categorical data differences were carried out by chi-square analysis. For every patient, we checked the adherence to the protocol during the study; for patients not completely meeting the protocol standards, a note was written on the CRF. We assessed and reported total adherence rate to our protocol in order to analyse data on the intention-to-treat (ITT) and on the per-protocol (PP) population. Primary analysis was carried out on the ITT population. Significance level was established to be <0.05. Analyses were carried out with the ITT if not otherwise specified. Statistical data analysis was performed using the SPSS 26.0 package (SPSS Inc, USA).

## RESULTS

From May 2014 to February 2019, 474 consecutive patients were recruited, 27 of them were excluded, according to the study criteria. 429 patients were randomised into three parallel groups (Tab. 1 and Fig. 2).

**TABLE 1.** Patients’ characteristics stratified into control group, IVC ultrasound (IVCUS) group and Passive-Leg-Raising Test (PLRT) group. Continuous measurements were presented as mean (SD); categorical variables were reported as number (percentages).

|  | **Control**<br>**group**<br>**(n=149)** | **IVCUS**<br>**group**<br>**(n=132)** | **PLRT group**<br>**(n=148)** |
| --- | --- | --- | --- |
| **Age (y), (mean, SD)** | 58.7 (18.0) | 55.9 (18.3) | 57.2 (18.4) |
| **Weight (Kg), (mean, SD)** | 76.3 (17.2) | 76.7 (16.7) | 77.8 (15.1) |
| **ASA status** |  |  |  |
| I – II, *n* (%) | 119 (80%) | 114 (86%) | 130 (88%) |
| III, *n* (%) | 30 (20%) | 18 (14%) | 18 (12%) |
| **Anti-hypertensive therapy, *n* (%)** | 53 (36%) | 37 (28%) | 38 (26%) |
| B-blockers, *n* (%) | 44 (30%) | 19 (14%) | 21 (14%) |
| ACE-inhibitors, *n* (%) | 17 (11%) | 16 (12%) | 14 (9%) |
| Other, *n* (%) | 15 (10%) | 16 (12%) | 15 (10%) |
| **Psychotherapy, *n* (%)** | 4 (3%) | 7 (5%) | 11 (7%) |
| SSRI, *n* (%) | 3 (2%) | 4 (3%) | 2 (1%) |
| iMAO, *n* (%) | - | - | - |
| Other, *n* (%) | 2 (1%) | 3 (2%) | 9 (6%) |

**FIGURE 2:**
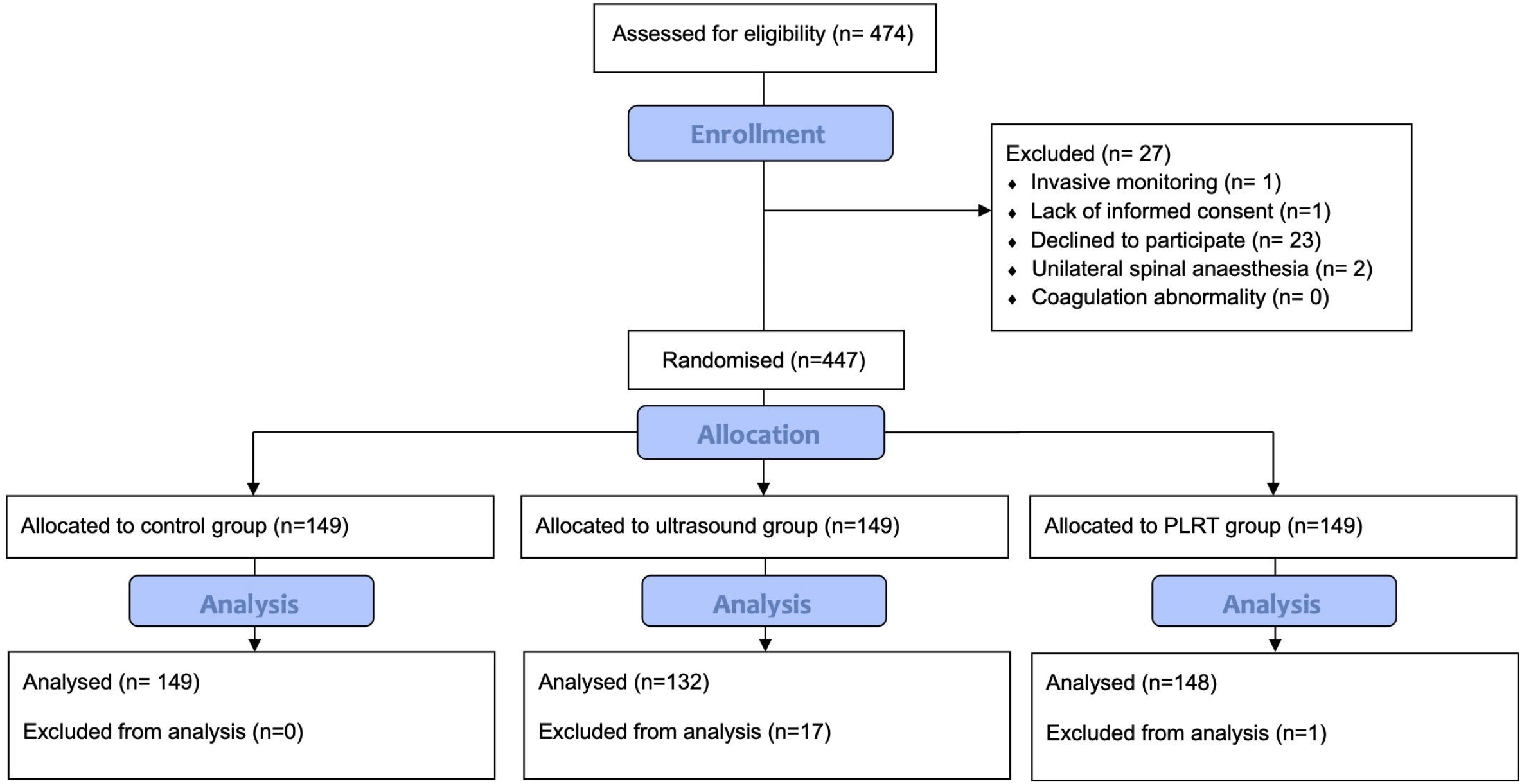
study flow chart. Structure of patients’ enrolment, allocation and analysis according to study design.

The average amount of fluids administered to the patient before SA was 141 ml (±135 ml) for the control group, 336 ml (±314 ml) for the IVCUS group and 168 ml (±237 ml) for the PLRT group (*P*<0.001). Patients who received a pre-anaesthesia volume filling greater than 500 ml were 8 in control group, 58 in IVCUS group and 25 in PLRT group (Fig. 3, Tab. 2 and 3).

**FIGURE 3:**
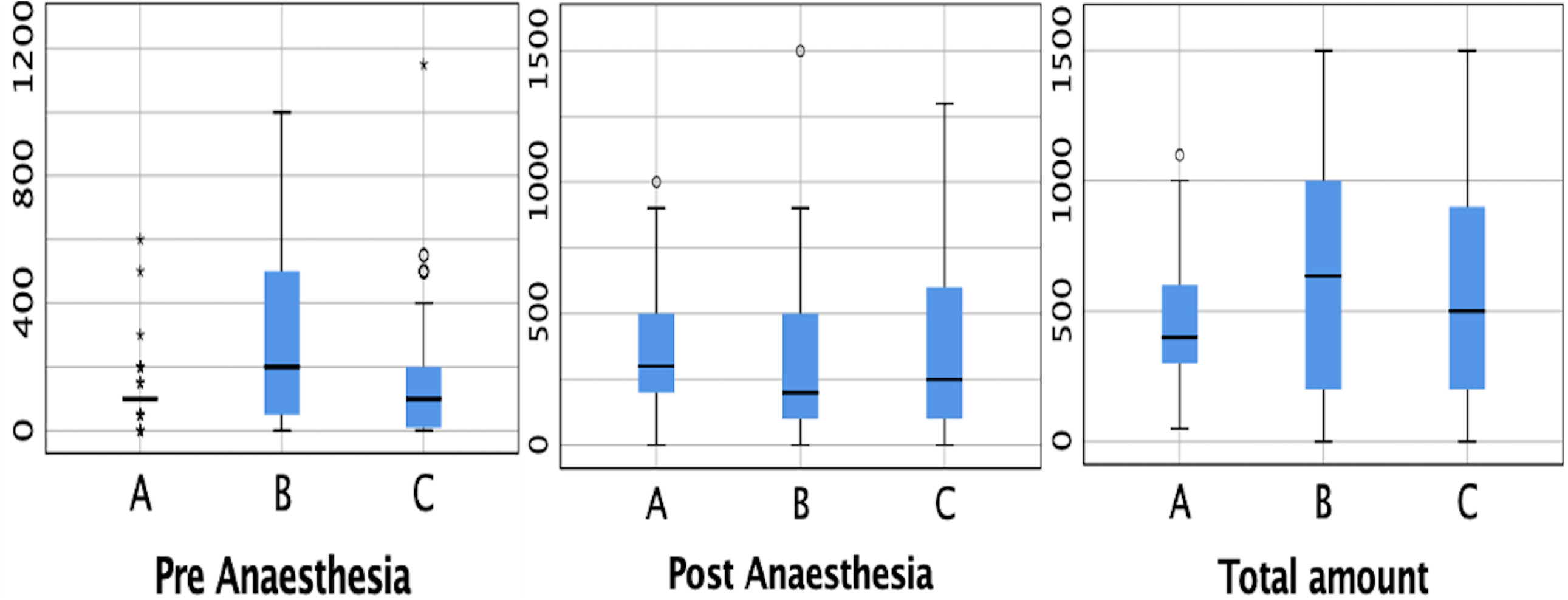
fluid administration. Boxplot representing the crystalloids administration in control group (A), in the IVCUS group (B) and in the PLRT group (C). Data of the pre-anaesthesia phase, postanaesthesia phase and globally (pre- and postanaesthesia phase) are reported.

**TABLE 2.** Hemodynamic patients’ characteristics between control group and IVCUS group, at the anaesthesia, the post-anaesthesia and globally (*pre-* and *post-anaesthesia*).

|  | **Control group**<br>**(n=149)** | **IVCUS**<br>**group**<br>**(n=132)** | **P value** |
| --- | --- | --- | --- |
| **At the anaesthesia** |  |  |  |
| Systolic AP (mmHg), (mean, SD) | 141 (21) | 140 (22) | 0.286 |
| Diastolic AP (mmHg), (mean, SD) | 77 (12) | 77 (10) | 0.510 |
| Heart Rate (mmHg), (mean, SD) | 74 (15) | 71 (11) | 0.096 |
| SpO<sub>2</sub>(%),(mean SD) | 97 (2) | 97 (2) | 0.169 |
| Fluid amount (ml), (mean, SD) | 141 (135) | 336 (314) | <0.001* |
| • NaCl 0.9%, n (%) | 2 (1%) | 2 (2%) | - |
| • Ringerfundine, n (%) | 147 (99%) | 130 (98%) | - |
| Minutes until anaesthesia, (mean, SD) | 22 (15) | 25 (13) | <0.095 |
| **Postanaesthesia** |  |  |  |
| Hypotension, n (%) | 68 (46%) | 46 (35%) | 0.047* |
| Additional fluid amount (ml), (mean, SD) | 312 (424) | 257 (266) | 0.117 |
| **Amine use** |  |  |  |
| • Ephedrine, n (%) | 48 (32%) | 18 (14%) | - |
| • Neosyneprine, n (%) | 5 (3%) | 2 (2%) | - |
| • Atropine, n (%) | 2 (1%) | 0 (0%) | - |
| Symptoms, n (%) | 10 (7%) | 6 (5%) | - |
| **Globally** |  |  |  |
| Total fluid amount (ml), (mean, SD) | 453 (458) | 593 (369) | 0.015* |
| SAE, n (%) | 1 (1%) | 0 (0%) | - |

Continuous measurements were presented as mean, SD; categorical variables were reported as number (percentages). Significance was established at $P<0.05$ (\*).

**TABLE 3.** Hemodynamic patients’ characteristics between control group and PLRT group, at the anaesthesia, the post-anaesthesia and globally (*pre-* and *post-anaesthesia*).

|  | **Control**<br>**group**<br>**(n=149)** | **PLRT group**<br>**(n=148)** | **P value** |
| --- | --- | --- | --- |
| **At the anaesthesia** |  |  |  |
| Systolic AP (mmHg), (mean, SD) | 141 (21) | 142 (25) | 0.28 |
| Diastolic AP (mmHg), (mean, SD) | 77 (12) | 79 (11) | 0.51 |
| Heart Rate (mmHg), (mean, SD) | 74 (15) | 70 (11) | 0.09 |
| SpO<sub>2</sub>(%), (mean, SD) | 97 (2) | 97 (2) | 0.16 |
| Fluid amount (ml), (mean, SD) | 141 (135) | 168 (237) | 0.77 |
| • NaCl 0.9%, *n* (%) | 2 (1%) | 2 (1%) | - |
| • Ringerfundine, *n* (%) | 147 (99%) | 146 (99%) | - |
| Minutes until anaesthesia, (mean, SD) | 22 (15) | 29 (17) | 0.07 |
| **Postanaesthesia** |  |  |  |
| Hypotension, *n* (%) | 68 (46%) | 65 (44%) | 0.15 |
| Additional fluid amount (ml), (mean, SD) | 312 (424) | 342 (321) | 0.75 |
| **Amine use** |  |  |  |
| • Ephedrine, *n* (%) | 48 (32%) | 29 (20%) | - |
| • Neosyneprine, *n* (%) | 5 (3%) | 2 (1%) | - |
| Atropine use, *n* (%) | 2 (1%) | 0 (0%) | - |
| Symptoms, *n* (%) | 10 (7%) | 8 (5%) | - |
| **Globally** |  |  |  |
| Total fluid amount (ml), (mean, SD) | 453 (458) | 511 (368) | 0.02* |
| Serious Adverse Events, *n* (%) | 1 (1%) | 0 (0%) | - |

Continuous measurements were presented as mean, SD; categorical variables were reported as number (percentages). Significance was established

### Primary outcome

After performing SA, the number of patients affected by arterial hypotension was 68 (46%) in the control group, 46 (35%) in the IVCUS group and 65 (44%) in the PLRT group. Concerning hypotension rates between groups, the IVCUS group compared with control group showed a reduction of 11% (95% CI -1 to -24%, *P*=0.047); the PLRT group compared with control group had a reduction around 2% (95% CI -3 to -5%, *P*=0.428).

### Secondary outcomes

The amount of fluids administered postanaesthesia was 312 ml (±424 ml), 257 ml (±266 ml) and 342 ml (±321 ml) for the control, IVCUS and PLRT group respectively without a significant difference (*P*=0.117). Globally the total amount of fluids administered was 453 ml (±458 ml), 593 ml (±369 ml) and 511 ml (±368 ml) for the control, IVCUS and PLRT group respectively, with a greater administration in the IVCUS group (*P*=0.015). No patient developed any adverse event, especially sign of fluid overload.

Concerning the use of hemodynamically active drugs, ephedrine was used in 32% of patients in the control group, in 14% of IVCUS patients and in 20% of patients in the PLRT group (Tab. 2 and 3).

The total study time for each patient was 46 min (±27 min) for the control group, 48 min (±10 min) for the IVCUS group and 40 minutes (±13 min) for the PLRT group; comparing global anaesthesia average time between IVCUS and control group, no difference was found (*P*=0.35, Fig. 4).

**FIGURE 4:**
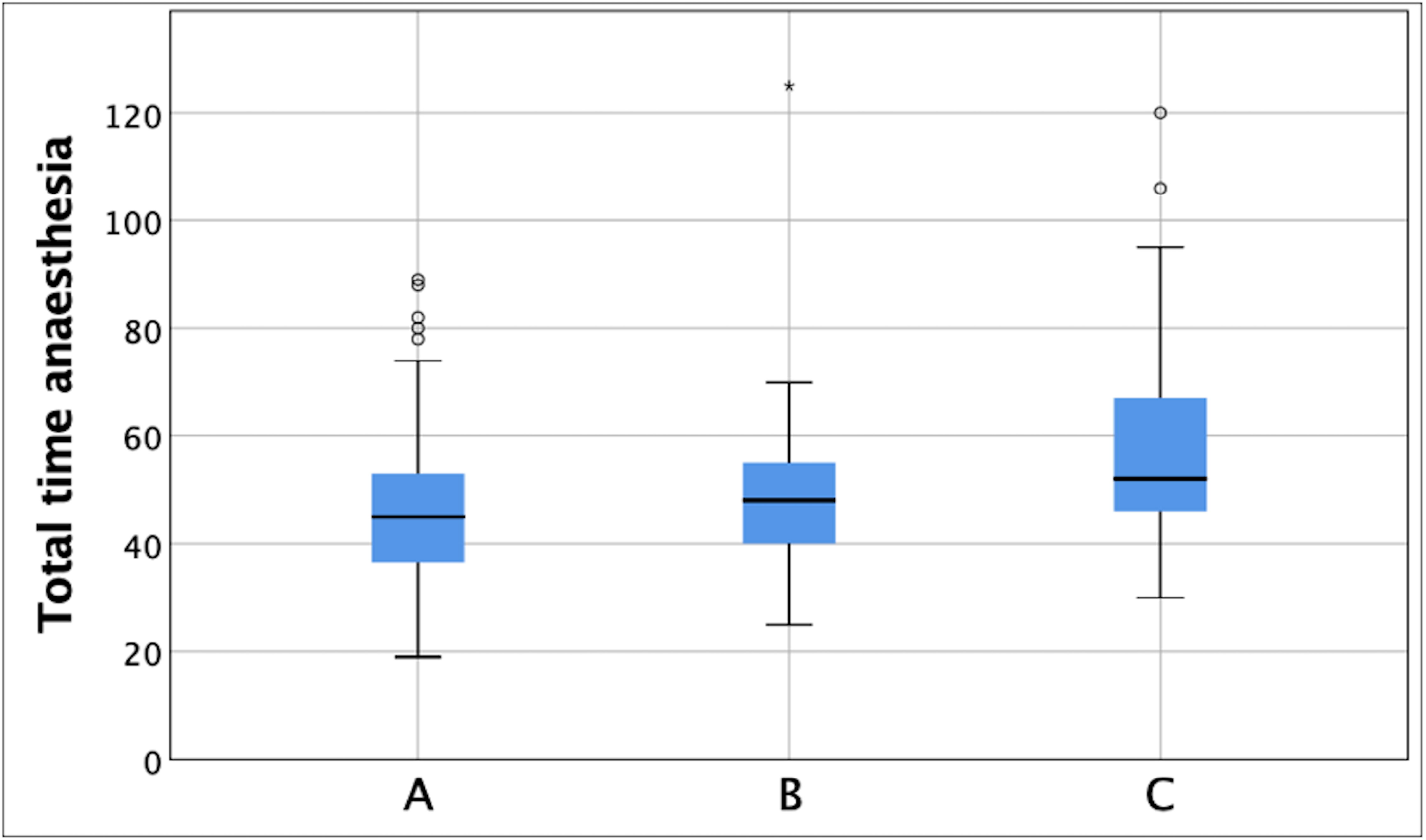
total time stratification. Representative boxplot comparing the total time required to complete anaesthesia in the control group (A), in the IVCUS group (B) and in the PLRT group (C). Data of the pre-anaesthesia phase, postanaesthesia phase and globally (pre- and postanaesthesia phase) are reported.

### Adherence

The adherence to the protocol was adequate in 96% of cases; 100% in the control group, 89% in the echo-group, 99.4% in the PLRT group (Fig. 2). Most of patients excluded from the protocol were in the IVCUS group due to the non-feasibility of IVCUS measurement for intrinsic patient’s characteristic (e.g. BMI, windows echogenicity). One single patient in PLRT group was considered out of adherence since he was sedated.

## DISCUSSION

SA is a common anaesthetic procedure burdened by arterial hypotension [2, 5, 9]. Usually, this has mild consequences in healthy patients, often presenting transitory dizziness, nausea or vomiting [7, 8], but it can lead to more serious consequences in groups of patients with pulmonary, cardiac and cerebrovascular diseases [4, 9]. As previously reported in the literature, the rate of arterial hypotension is around 50% [20]; in the standard group of this trial we reported a prevalence of 46%, confirming that this complication remains extremely frequent. The development of a method able to reduce this hurdle could be a relevant strategy to potentially reduce post anaesthesia morbidity. To our knowledge, currently no previous trial compared post SA hypotension rate using fluid administration by IVC-ultrasound or by PLRT in non-obstetrical patients.

The pre-clinical question was to assess whether IVCUS was a valid tool to decrease the rate of arterial hypotension by identifying fluid responsive patients in spontaneous breathing. Main result of this study was marked hypotension rate drop (−11%) between the IVCUS and the control group, which is in line with what was previously found [21]. If compared to the values reported by the literature (at least 15%), this reduction rate could appear as relatively small. However, this result concerns an extremely frequent pathology, such as post induction hypotension (prevalence of 50%) [20], with an important impact, from 1/3 to half, on the examined population. Evaluating this aspect, an 11% reduction appears extremely promising. A slight decrease in the hypotension rate was registered comparing the PLRT group with standard treatment.

These methods lead to an increase in amount of crystalloids administered prior to anaesthesia, significant in the IVCUS group compared with control group. This could potentially lead to fluid overload. However, this greater fluid amount was comparable to the standard group: after each fluid bolus an IVC-ultrasound check was performed in order to verify the patient’s volaemic status, carefully avoiding any fluid overload.

Regarding the PLRT group, the pre-anaesthesia fluid amount was not significantly different compared with control group.

According to other authors, a greater pre-anaesthesia fluid administration is responsible for arterial hypotension rate drop [26 - 28]. A key element is that this filling was not carried out indiscriminately but was defined on the basis of two methods that potentially were able to identify patients who can benefit from this filling and to identify patients that do not need further fluid, avoiding any unsafe fluid overload. Assessing the impact of these two non-invasive methods in reducing post SA arterial hypotension, such as PLRT and IVCUS, our trial showed positive results towards the use of IVC-ultrasound to identify spontaneous-breathing patients needing fluid administration prior to SA.

Previous study showed that IVCUS was unable to predict the risk of arterial hypotension after SA [29]. It’s important to highlight that our trial did not analyse this aspect; we focused on the *IVCUS analysis* as a useful procedure in identifying patients who could benefit of a fluid refilling. Further, these techniques reduce the use of vasopressors compared with standard treatment, especially in the IVCUS group (saving up to 60% of global amine usage), potentially eluding any other iatrogenic complications related to drug consumption.

Our trial had some limitations. First, it was not possible to blind patients about group allocation; to overcome this limitation we chose to clearly define all the controlling factors required to describe patient as “hypotensive patient”, to avoid any subjective definition [20]. Moreover, to reduce further bias in this setting, the identification of the patient’s criteria of arterial hypotension was determined by the software and not by the physician in charge, according to these pre-determined limits [20]. Second, ultrasounds could be not feasible in a certain number of patients due to poor ultrasound windows, which explains the 12% of dropout rate we recorded in the IVCUS group; this data was probably underestimated in our trial for a low patient ratio with uncomfortable ultrasound features (e.g. obesity, COPD, etc.) and for a good anaesthesiologist expertise in ultrasound performance.

Finally, the sample studied was generalizable simply to a part of the whole population, as we included only ASA I to III patients with low cardiovascular risk. ASA IV and high cardiovascular risk patients were excluded to avoid any exposition to risk of arterial hypotension or fluid overload to any patient at risk. In this context, it is remarkable to remember that some authors showed that SA is often performed instead of general anaesthesia in high cardiovascular risk patient needing hip surgery. An intriguingly future trial should include patients at higher cardiovascular risk to test robustness of our results.

In conclusion, incidence of arterial hypotension after SA is extremely frequent. This study provides encouraging evidence that IVCUS combined with a fluid protocol may decrease the incidence of arterial hypotension after SA.

## Data Availability

All data are available under specific request

## ACKNOWLEDGEMENTS RELATING TO THIS ARTICLE

Assistance with the study: We would like to thank Alessandro Spila, Ciesse Sistemi Bologna (I) for his assistance with the study.

Financial support and sponsorship: none.

Conflicts of interest: none.

Presentation: preliminary data for this study were presented at the SSICM annual meeting on the 19^th^ of September 2019 in Lausanne, Switzerland, and at the national congress “Réanimation 2020 SRLF” on the 7^th^ of February 2020, in Paris, France.

